# Assessment of COVID-19 intervention strategies in the Nordic countries using genomic epidemiology

**DOI:** 10.1101/2021.09.04.21263123

**Authors:** Sebastian Duchene, Leo Featherstone, Birgitte Freiesleben de Blasio, Edward C. Holmes, Jon Bohlin, John H.-O. Pettersson

## Abstract

The Nordic countries, defined here as Norway, Sweden, Denmark, Finland and Iceland, are known for their comparable demographics and political systems. Since these countries implemented different COVID-19 intervention strategies, they provide a natural laboratory for examining how COVID-19 policies and mitigation strategies affected the propagation, evolution and spread of the SARS-CoV-2 virus. We explored how the duration, the size and number of transmission clusters, defined as country-specific monophyletic groups in a SARS-CoV-2 phylogenetic tree, differed between the Nordic countries. We found that Sweden had the largest number of COVID-19 transmission clusters followed by Denmark, Norway, Finland and Iceland. Moreover, Sweden and Denmark had the largest, and most enduring, transmission clusters followed by Norway, Finland and Iceland. In addition, there was a significant positive association between transmission cluster size and duration, suggesting that the size of transmission clusters could be reduced by rapid and effective contact tracing. Thus, these data indicate that to reduce the general burden of COVID-19 there should be a focus on limiting dense gatherings and their subsequent contacts to keep the number, size and duration of transmission clusters to a minimum. Our results further suggest that although geographical connectivity, population density and openness influence the spread and the size of SARS-CoV-2 transmission clusters, country-specific intervention strategies had the largest single impact.

## Introduction

The Nordic countries of northern Europe - Norway, Denmark, Iceland, Finland and Sweden - are known for their similar demographics and political systems. However, during the first year of the COVID-19 pandemic, and prior to widespread vaccination roll-out, these countries adopted varied policy responses and intervention measures (1). While Sweden initially implemented a more lenient mitigation strategy, Norway, Denmark and Finland applied inhibitory non-pharmaceutical interventions that included a widespread population “lock-down”. Iceland, a small isolated island population, focused on contact tracing and testing (1). Because of their homogeneity, the Nordic countries therefore provide a natural laboratory to compare COVID-19 intervention strategies (2, 3). We recently analysed transmission clusters, defined as country-specific monophyletic groups derived from a phylogenetic analysis of the causative Severe Acute Respiratory Syndrome Coronavirus 2 (SARS-CoV-2), in the Nordic countries using a genomic data set covering the Nordic region (1). Here, we use the same transmission clusters to analyse the epidemiological implications of the different intervention strategies employed by the Nordic health authorities.

## Methods

Briefly, the data was generated in (1) by (i) downloading and aligning all available Nordic SARS-CoV-2 genomes from GISAID (www.gisaid.org; with 67,918 Nordic genomes) and the NextStrain global build as of 22nd March 2021 (3,437 global genomes). The data set was then subsampled ten times (referred to here as “replicates”) according to prevalence per country, resulting in alignments including between 15,297 and 15,616 SARS-CoV-2 genome sequences. For each alignment, a phylogenetic tree was computed with IQ-TREE v2.0.6 (4) scaled to time using LSD v.03 (5) under a strict molecular clock at a fixed rate of 1×10^−3^ nucleotide substitutions/site/year, and the GTR+Γ substitution model. Transmission clusters were defined as the monophyletic clustering of two or more sequences from the same country. The duration of a transmission cluster was defined as the time between the last and the first sample. The data set was converted to a weekly time-series format, calculated from the time to most recent common ancestor (TMRCA) from each transmission cluster. Analyses were carried out using generalised additive mixed-effects models (GAMM) employing restricted maximum likelihood (REML) (6). We used GAMM for the regression models with country as the explanatory variable and the following outcomes: number of transmission clusters, transmission cluster size (*i*.*e*. the number of samples in each transmission cluster) and transmission cluster duration. An additional GAMM model was included to assess the association between duration and transmission cluster size. This model included an interaction term between country and number of samples as the explanatory variable with duration as the outcome. All models, except that having the number of transmission clusters as an outcome, were also adjusted for the number of transmission clusters and TMRCA with respect to country using splines. Replicate number (1–10) by country (Norway, Sweden, Finland, Denmark and Iceland) was included as a hierarchical random effect in all models. Goodness-of-fit was determined using Akaike Information Criterion (AIC), R^2^ and model residuals conformance to normality. Regression analyses were performed using the GAMM4 package (6), and figures were created using the ggplot2 package (7). A description of the statistical estimation procedures and results are provided in the Supplementary Appendix 1.

## Results

We divided the data set into the number of transmission clusters per week. First, we compared the number of transmission clusters between the different countries (see Supplementary Figure S1 and the Methods section for more details regarding the regression models). This revealed that Sweden had significantly more transmissions clusters (p<0.001) than Denmark, who in turn had significantly more than Norway (p<0.001). The number of transmission clusters in Finland was comparable to Norway (p=0.878). The fewest number of transmission clusters were found in Iceland (p<0.012), but the smoothing spline did not fit the data properly (p=0.906) due to the low number of cases.

We next compared transmission cluster size *(i*.*e*. number of samples per cluster) between the different Nordic countries. These analyses suggest that transmission cluster size did not differ significantly between Sweden and Denmark (p=0.895). However, transmission clusters were significantly smaller (p<0.001) in Norway, followed by Finland (p<0.001), while Iceland’s model was not found to be significant (p=0.898). Figure 1 shows the log-transformed number of samples per transmission cluster (*i*.*e*. transmission cluster size) with respect to TMRCA, together with regression model lines, for each Nordic country and all replicates. During the autumn and winter months of 2020 (September through December) we see an increase in transmission cluster size, with the exception of Finland and Iceland (Figure 1).

**Figure 1.**
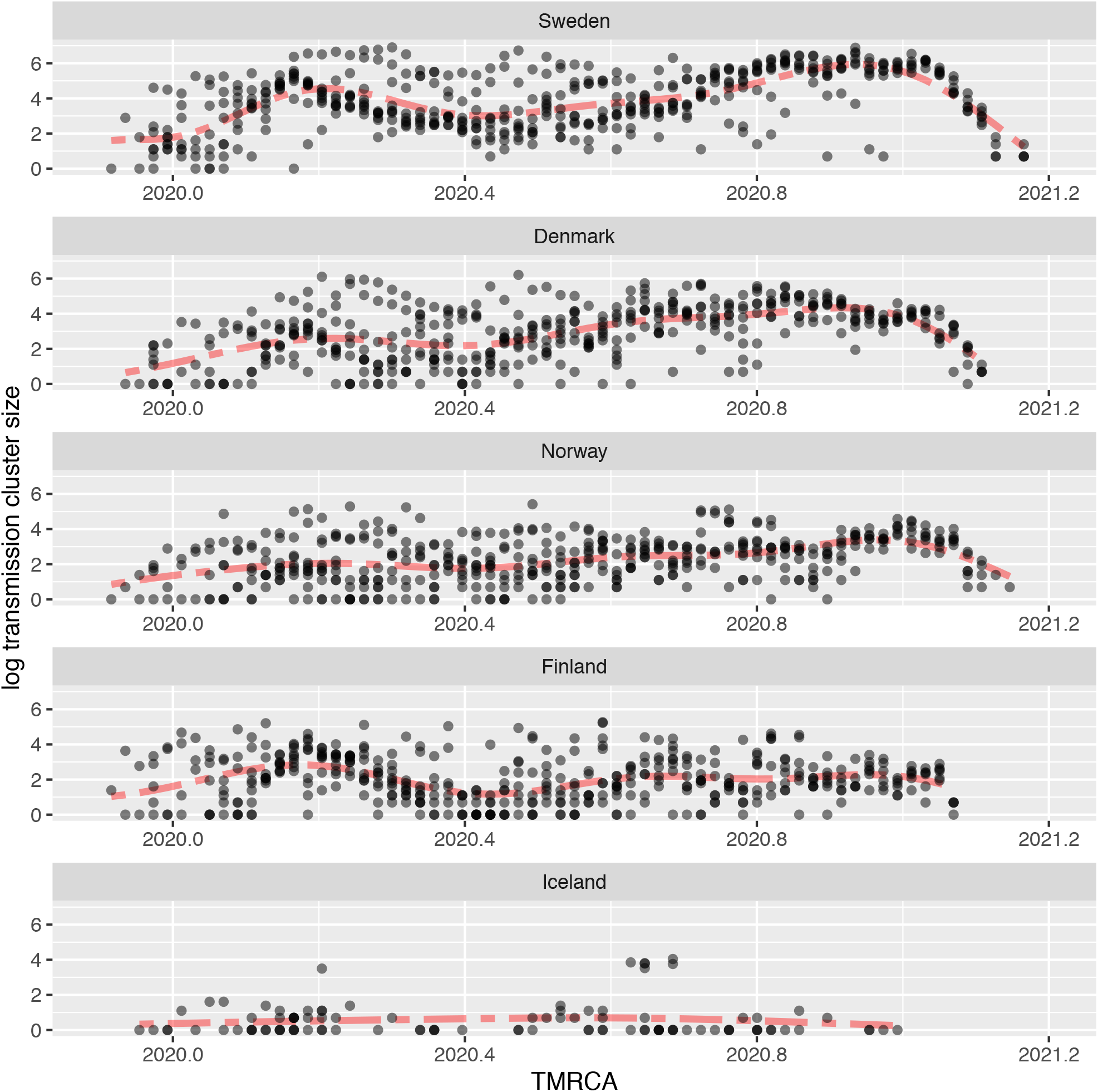
The (log transformed) size of transmission clusters (*i*.*e*. number of cases per transmission cluster, vertical axis) for replicates 1–10 with respect to the TMRCA (time to most recent common ancestor, horizontal axis) for the Nordic countries. The red dashed line represents the regression model averaged over all replicates.

We also compared transmission cluster duration between the Nordic countries. This did not significantly differ between Sweden and Denmark (p=0.269) who, in turn, had significantly more enduring transmission clusters than the other Nordic countries (p<0.002). There was no significant difference between Norway and Finland (p=0.503), while Iceland’s model was again not significant (p=0.887) due to the low number of cases. In Figure 2 we see that the duration of the transmission clusters drops when the winter months approach the end of 2020. A significant positive association (p<0.001) between transmission cluster size and duration was established suggesting that the duration of a transmission cluster correlates with its size.

**Figure 2.**
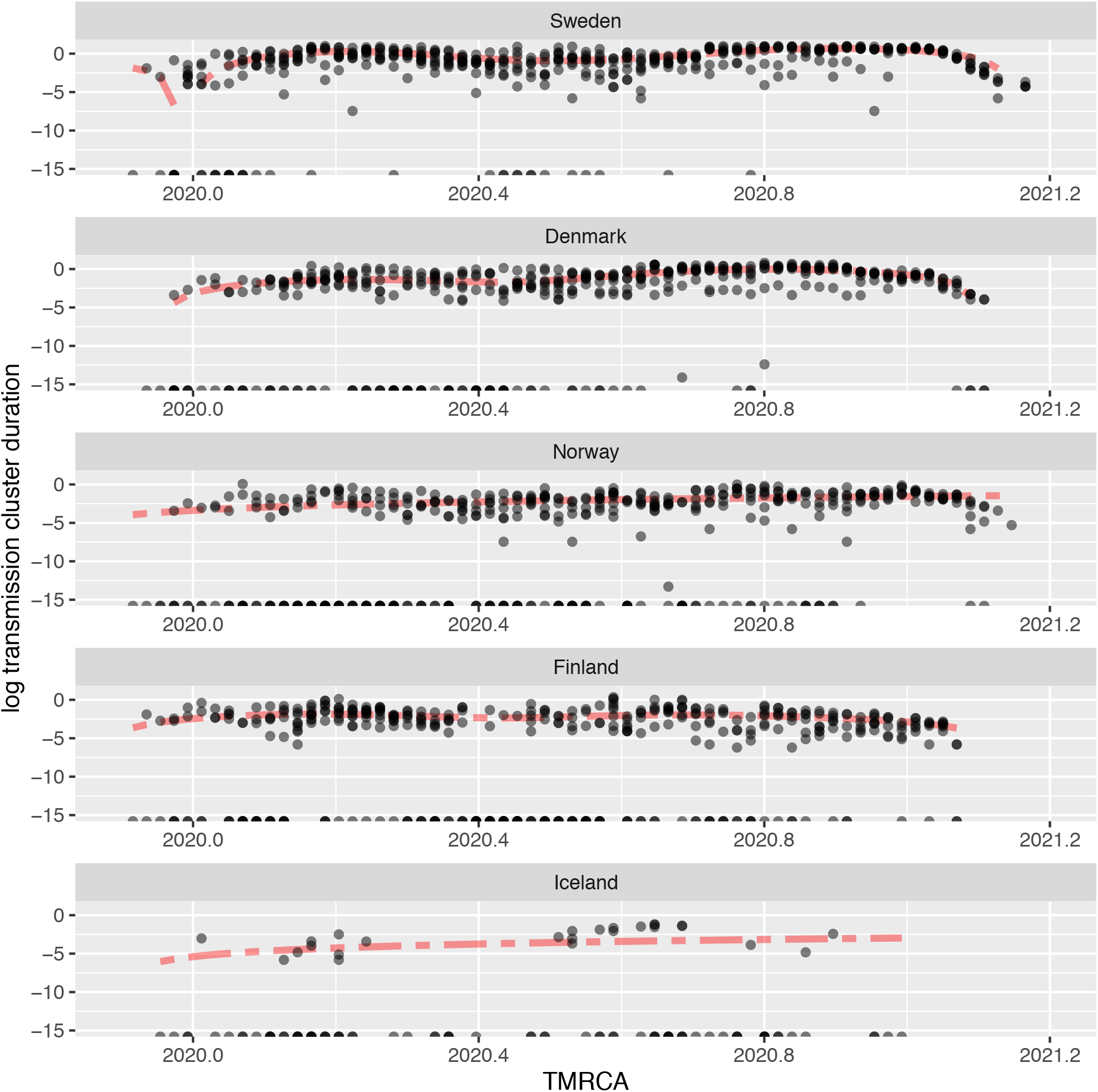
The (log transformed) transmission cluster duration (difference between last and first sample, vertical axis) for replicates 1–10 with respect to TMRCA (time to most recent common ancestor, horizontal axis) for the Nordic countries. The red dashed line represents the regression model averaged over all replicates.

## Discussion

We examined how the number of transmission clusters and their size and duration, differed among the Nordic countries. The presence of many samples within a transmission cluster indicate that one common source is responsible for many subsequent cases, while a low number points to fewer secondary infections. Accordingly, Sweden had the largest number of transmission clusters and, together with Denmark, also the most enduring. Moreover, our results also point to an increase of COVID-19 transmission clusters during winter, which could be due to the cold winter climate leading to increased indoor-based activities and thus increased contact frequencies for transmission (8). Alternatively, the increased spread of SARS-CoV-2 during the winter months could result from more relaxed COVID-19 intervention measures during the autumn (1). Notably, transmission cluster size decreased towards the end of 2020 in concordance with an increase in the governmental stringency index for the Nordic countries (1). Hence, while climate and change of season influence behaviour and contact frequency, our analyses suggest that the adopted intervention measures also effectively reduced virus transmission.

The largest transmission clusters were found in Sweden and Denmark, while the size of the transmission clusters in Norway and Finland were significantly smaller and more similar. Iceland differed in general from the other Nordic countries with substantially fewer COVID-19 cases, which can most likely be ascribed to the country’s small population and isolated location. Interestingly, although Sweden had more transmission clusters (Supplementary Figure S1), their duration and size was not significantly different from the clusters obtained from Denmark. Although we have tried to adjust for bias in sequencing intensity by using sub-sampled SARS-CoV-2 sequence data from each country in the regression models (1), some bias may still persist due to country-specific strategies for selecting cases for sequencing. In particular, Swedish transmission clusters could be underestimated in terms of both size and duration. Alternatively, the more stringent intervention policies implemented in Denmark, as compared to Sweden, could have reduced the total number of infections but allowed the transmission clusters to endure. Indeed, we found a significant association between transmission cluster size and duration. Finland and Iceland had the most short-lived transmission clusters, with Iceland’s significantly shorter than Finland’s.

During the second half of 2020, Sweden adopted stricter COVID-19 intervention policies, more on par with its neighbouring countries Norway and Finland, but the number of cases remained high (1). This may point to some inertia regarding COVID-19 policies, such that the attitudes in the population and consequences from early COVID-19 intervention strategies persisted for some time after the new measures were introduced. As Norway and Finland are similar to Sweden in terms of demographics, location, climate and governance, our results demonstrate the effects of the variable COVID-19 control measures adopted. Norway, Finland and Denmark experienced a similar burden, while significantly more cases and transmission clusters were noted for Denmark. The increased number of cases and transmission clusters observed in Denmark, compared to Norway and Finland, could in part be due to far more intensive SARS-CoV-2 sequencing, although it may also be a consequence of Denmark’s higher population density and placement on the European continent. We observed a similar pattern in our previous study (1), in which both Denmark and Sweden had the most importation events (as well as the most exportation events). The duration of transmission clusters in Norway and Finland, that implemented similar restriction measures to Denmark, was significantly shorter compared to Denmark and Sweden. The Norwegian and Finnish strategies have focused on municipality-based, rigorous contact tracing, isolation and quarantining with the aim to clamp down transmission.

Our results suggest that reducing transmission cluster duration through effective contact tracing could also reduce transmission cluster size. The shorter duration of transmission clusters in Norway and Finland may reflect the effectiveness of the strategies deployed in those countries. Importantly, a growing number of transmission clusters would likely imply an increase in the genetic diversity of SARS-CoV-2, while the presence of transmission clusters of extended length better enable the virus to evolve.

The SARS-CoV-2 Alpha variant of concern (Pangolin lineage B.1.1.7) was first discovered in the United Kingdom and was associated with an increase in infectivity (9). Interestingly, we did not see any dramatic change in transmission cluster size or duration associated with the introduction of the Alpha variant. Indeed, with the exception of Iceland, both the duration and number of cases changed markedly in all countries from mid-2020 onwards, with cases dropping markedly at the end of 2020 (Figures 1 and 2) and not contemporaneous with the rise of the Alpha variant. The lack of any clear resurgence associated with the emergence of the Alpha variant likely reflects the impact of the COVID-19 control policies in place (1), again emphasizing their importance in controlling the pandemic.

The strategies implemented by Norway, Finland and Iceland, appear to have been more successful in reducing both the number and the size of their transmission clusters than those in Denmark and Sweden. As Sweden is a geographical neighbour to Norway and Finland (Denmark is located on the European continent neighbouring Germany) our findings highlight the importance of local measures in controlling the spread of SARS-CoV-2. However, it is unclear to what extent geographical location or strict COVID-19 strategies are responsible for the lower burden of COVID-19 in Norway, Finland and Iceland compared to Sweden and Denmark. Controlling outbreaks in Denmark may be more challenging as it is a relatively small country, located on the European continent, with a comparatively high population density. In contrast, the pattern in Iceland may reflect the fact that it is an island with a substantially smaller population located far away from other countries. Our findings from Iceland resembled those of other small island nations (10).

## Conclusions

Using a genomic epidemiology-based approach we found that transmission cluster size and duration differed markedly among the Nordic countries. Considering the homogeneous populations and the similar political systems in these countries, our findings clearly demonstrate that COVID-19 intervention policies can have profound influence on how epidemics evolve at the scale of individual countries. In particular, as Norway, Sweden and Finland are neighbouring countries with similar demographics, political systems and climate, the data presented here highlight the influence of intervention strategies in controlling the spread of COVID-19. Interestingly, Denmark implemented more similar COVID-19 intervention strategies to Norway and Finland, but did not reduce the spread of COVID-19 to same extent as other Nordic countries which may reflect its location on the European continent and relatively high population density. Hence, our results suggest that disease intervention strategies should be adapted to the specific geographic and demographic factors of each country to more effectively reduce the transmission and evolution of SARS-CoV-2.

## Data Availability

All SARS-CoV-2 genomes are available via the GISAID platform (www.gisaid.org).

http://www.gisaid.org

## Appendix

Supplementary File 1: Appendix 1. A pdf-file containing regression model details and statistics.

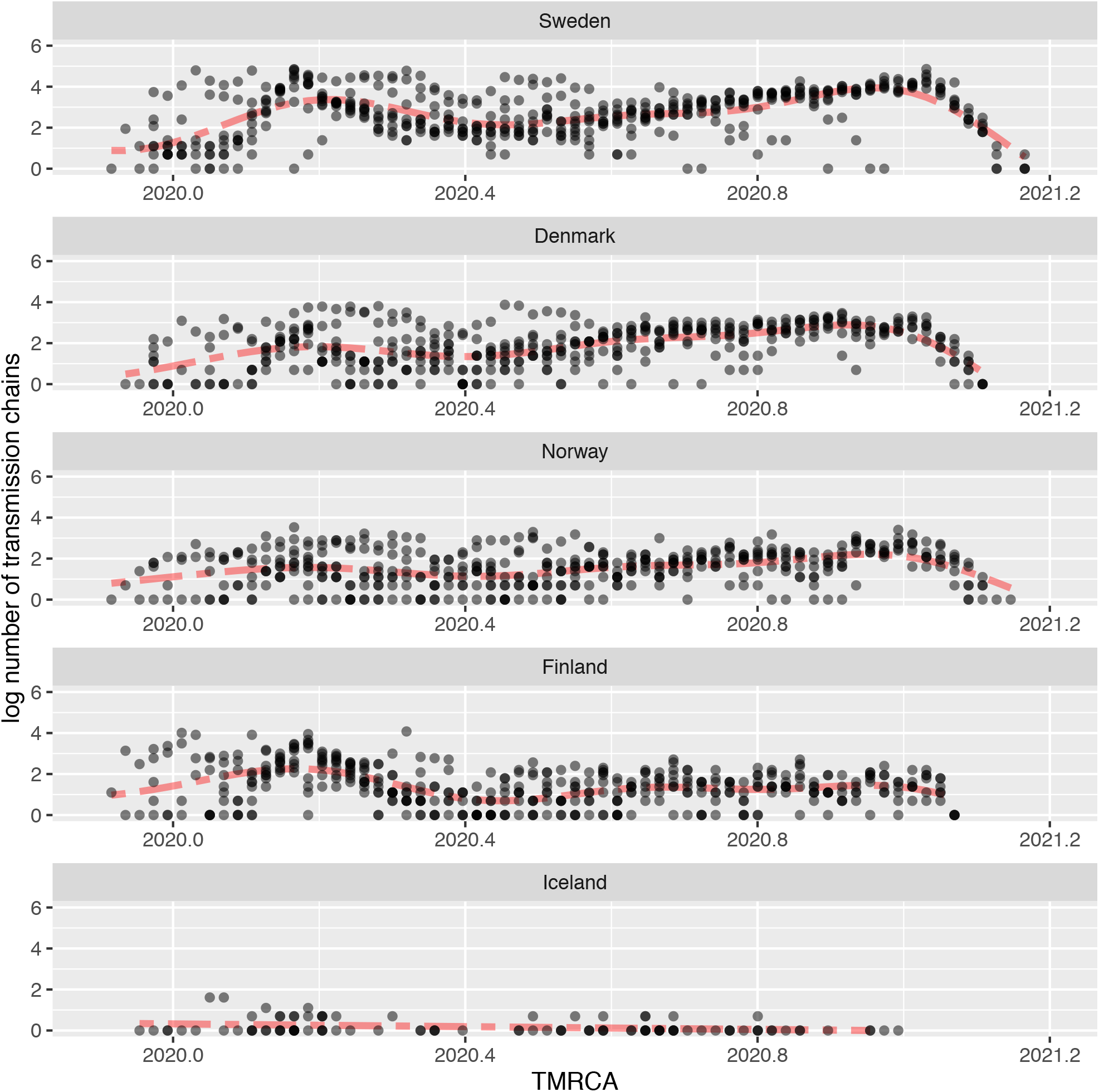

**Supplementary File 2**. Figure S1. The (log transformed) number of transmission clusters (vertical axis) for replicates 1–10 with respect to the TMRCA (time to most recent common ancestor, horizontal axis) for the Nordic countries. The red dashed line represents the regression model averaged over all replicates.

