## Supplementary file 1 for "Assessment of COVID-19 intervention strategies in the Nordic countries using genomic epidemiology"

Model: Number of transmission clusters

```
mod.gam<-gamm4(num_chains ~ country  + s(tmrca, by=country), random =~
(1|replicate/country), data=myfrm2)

Sweden compared against the other Nordic countries:

Scaled residuals:
    Min      1Q  Median      3Q     Max
-4.9084 -0.3663 -0.0786  0.2173  7.9180

Parametric coefficients:
               Estimate Std. Error t value Pr(>|t|)
(Intercept)      27.485      1.273   21.59   <2e-16 ***
countryDenmark  -16.959      1.265  -13.40   <2e-16 ***
countryNorway   -20.600      1.254  -16.42   <2e-16 ***
countryFinland  -20.785      1.260  -16.50   <2e-16 ***
countryIceland  -24.878      1.723  -14.44   <2e-16 ***
---
Signif. codes:  0 ‘***’ 0.001 ‘**’ 0.01 ‘*’ 0.05 ‘.’ 0.1 ‘ ’ 1

Approximate significance of smooth terms:
                          edf Ref.df      F  p-value
s(tmrca):countrySweden  7.640  7.640 84.711  < 2e-16 ***
s(tmrca):countryDenmark 5.884  5.884  7.996 2.35e-08 ***
s(tmrca):countryNorway  1.000  1.000  7.832  0.00518 **
s(tmrca):countryFinland 4.672  4.672  6.118 1.40e-05 ***
s(tmrca):countryIceland 1.000  1.000  0.014  0.90572
---
Signif. codes:  0 ‘***’ 0.001 ‘**’ 0.01 ‘*’ 0.05 ‘.’ 0.1 ‘ ’ 1

R-sq.(adj) =  0.433
lmer.REML =  15236  Scale est. = 144.97    n = 1939

Norway compared against the other Nordic countries:
Parametric coefficients:
               Estimate Std. Error t value Pr(>|t|)
(Intercept)      6.8854     1.2328   5.585 2.67e-08 ***
countryDenmark   3.6410     1.2478   2.918  0.00356 **
countrySweden   20.5996     1.2544  16.422  < 2e-16 ***
countryFinland  -0.1851     1.2231  -0.151  0.87970
countryIceland  -4.2787     1.7106  -2.501  0.01246 *
---
Signif. codes:  0 ‘***’ 0.001 ‘**’ 0.01 ‘*’ 0.05 ‘.’ 0.1 ‘ ’ 1

Random effects:
 Groups            Name                    Variance  Std.Dev.
 country:replicate (Intercept)             4.139e+00   2.035
 replicate         (Intercept)             7.804e+00   2.793
```

```
 Xr.3              s(tmrca):countryIceland 0.000e+00   0.000
 Xr.2              s(tmrca):countryFinland 2.446e+03  49.454
 Xr.1              s(tmrca):countryNorway  0.000e+00   0.000
 Xr.0              s(tmrca):countryDenmark 5.086e+03  71.319
 Xr                s(tmrca):countrySweden  1.956e+05 442.237
 Residual                                  1.450e+02  12.040
Number of obs: 1939, groups:
country:replicate, 45; replicate, 10; Xr.3, 8; Xr.2, 8; Xr.1, 8; Xr.0, 8;
Xr, 8
```

<u>Model: Size of transmission clusters</u>

```
mod.gam<-gamm4(num_samples ~ country  + s(num_chains, by=country) +
s(tmrca, by=country), random =~ (1|replicate/country), data=myfrm2)

Sweden compared against the other Nordic countries:

Scaled residuals:
    Min      1Q  Median      3Q     Max
-6.0944 -0.2106 -0.0592  0.0573 13.1772

Parametric coefficients:
               Estimate Std. Error t value Pr(>|t|)
(Intercept)      68.379      4.084  16.742  < 2e-16 ***
countryDenmark    0.993      7.499   0.132    0.895
countryNorway   -29.154      5.870  -4.967 7.42e-07 ***
countryFinland  -40.460      5.721  -7.073 2.12e-12 ***
countryIceland  -42.802    102.724  -0.417    0.677
---
Signif. codes:  0 ‘***’ 0.001 ‘**’ 0.01 ‘*’ 0.05 ‘.’ 0.1 ‘ ’ 1

Approximate significance of smooth terms:
                              edf Ref.df       F  p-value
s(num_chains):countrySweden  8.159  8.159 203.562  < 2e-16 ***
s(num_chains):countryDenmark 2.584  2.584 116.680  < 2e-16 ***
s(num_chains):countryNorway  1.000  1.000  40.168 2.88e-10 ***
s(num_chains):countryFinland 1.765  1.765  18.955 1.86e-05 ***
s(num_chains):countryIceland 1.000  1.000   0.041   0.8391
s(tmrca):countrySweden       6.779  6.779  35.778  < 2e-16 ***
s(tmrca):countryDenmark      3.772  3.772   3.278   0.0152 *
s(tmrca):countryNorway       1.000  1.000   0.257   0.6125
s(tmrca):countryFinland      1.056  1.056   0.764   0.4051
s(tmrca):countryIceland      1.000  1.000   0.079   0.7787
---
Signif. codes:  0 ‘***’ 0.001 ‘**’ 0.01 ‘*’ 0.05 ‘.’ 0.1 ‘ ’ 1

R-sq.(adj) =   0.71
lmer.REML =  21562  Scale est. = 4012.2    n = 1939
```

Norway compared against the other Nordic countries:

```
Parametric coefficients:
               Estimate Std. Error t value Pr(>|t|)    
(Intercept)      39.225      4.216   9.303  < 2e-16 ***
countryDenmark   30.153      7.579   3.978 7.20e-05 ***
countrySweden    29.154      5.870   4.967 7.42e-07 ***
countryFinland  -11.305      5.815  -1.944    0.052 .  
countryIceland  -13.657    106.108  -0.129    0.898    
---
Signif. codes:  0 ‘***’ 0.001 ‘**’ 0.01 ‘*’ 0.05 ‘.’ 0.1 ‘ ’ 1
```

```
Random effects:
 Groups            Name                          Variance  Std.Dev.
 country:replicate (Intercept)                   0.000e+00 0.000e+00
 replicate         (Intercept)                   0.000e+00 0.000e+00
 Xr.8              s(tmrca):countryIceland       1.235e-05 3.514e-03
 Xr.7              s(tmrca):countryFinland       4.459e+01 6.678e+00
 Xr.6              s(tmrca):countryNorway        4.949e-05 7.035e-03
 Xr.5              s(tmrca):countryDenmark       1.850e+04 1.360e+02
 Xr.4              s(tmrca):countrySweden        7.935e+05 8.908e+02
 Xr.3              s(num_chains):countryIceland 5.553e+04 2.356e+02
 Xr.2              s(num_chains):countryFinland 1.449e+04 1.204e+02
 Xr.1              s(num_chains):countryNorway  2.014e-02 1.419e-01
 Xr.0              s(num_chains):countryDenmark 1.168e+05 3.418e+02
 Xr                s(num_chains):countrySweden  1.355e+06 1.164e+03
 Residual                                        4.012e+03 6.334e+01
Number of obs: 1939, groups:
country:replicate, 45; replicate, 10; Xr.8, 8; Xr.7, 8; Xr.6, 8; Xr.5, 8;
Xr.4, 8; Xr.3, 8; Xr.2, 8; Xr.1, 8; Xr.0, 8; Xr, 8
```

Model: Duration of transmission clusters

```
mod.gam<-gamm4(duration ~ country + s(num_chains, by=country) +s(tmrca,
by=country), random =~ (1|replicate/country), data=myfrm2)
```

Sweden compared against the other Nordic countries:

```
Scaled residuals:
    Min      1Q  Median      3Q     Max 
-5.1852 -0.3946 -0.0926  0.2944  5.6194 
```

```
Parametric coefficients:
                Estimate Std. Error t value Pr(>|t|)    
(Intercept)     3.339e-01  4.315e-02   7.739 1.62e-14 ***
num_samples     2.448e-03  8.517e-05  28.748  < 2e-16 ***
countryDenmark -8.118e-02  7.338e-02  -1.106    0.269    
```

```
countryNorway  -2.347e-01  5.672e-02  -4.138 3.66e-05 ***
countryFinland -2.290e-01  4.512e-02  -5.075 4.25e-07 ***
countryIceland -3.382e-01  3.873e-01  -0.873    0.383
---
Signif. codes:  0 ‘***’ 0.001 ‘**’ 0.01 ‘*’ 0.05 ‘.’ 0.1 ‘ ’ 1

Approximate significance of smooth terms:
                              edf Ref.df       F  p-value
s(num_chains):countryNorway  1.319  1.319  42.182 1.31e-06 ***
s(num_chains):countryDenmark 3.227  3.227 109.838  < 2e-16 ***
s(num_chains):countrySweden  6.283  6.283 252.076  < 2e-16 ***
s(num_chains):countryFinland 2.418  2.418  23.896 2.37e-12 ***
s(num_chains):countryIceland 1.000  1.000   0.027  0.86897
s(tmrca):countryNorway       1.000  1.000   3.169  0.07519 .
s(tmrca):countryDenmark      7.512  7.512  25.699  < 2e-16 ***
s(tmrca):countrySweden       7.512  7.512  45.161  < 2e-16 ***
s(tmrca):countryFinland      2.933  2.933   4.400  0.00879 **
s(tmrca):countryIceland      1.000  1.000   0.020  0.88739
---
Signif. codes:  0 ‘***’ 0.001 ‘**’ 0.01 ‘*’ 0.05 ‘.’ 0.1 ‘ ’ 1

R-sq.(adj) =  0.757
lmer.REML = 825.22  Scale est. = 0.079783  n = 1939

Norway compared against the other Nordic countries:

Scaled residuals:
    Min      1Q  Median      3Q     Max
-5.5662 -0.3780 -0.0797  0.2989  6.5793

Parametric coefficients:
               Estimate Std. Error t value Pr(>|t|)
(Intercept)     0.20504    0.04724   4.340  1.5e-05 ***
countryDenmark  0.20165    0.06463   3.120  0.00184 **
countrySweden   0.30610    0.04944   6.191  7.3e-10 ***
countryFinland -0.03285    0.04900  -0.670  0.50271
countryIceland -0.13389    0.48195  -0.278  0.78120
---
Signif. codes:  0 ‘***’ 0.001 ‘**’ 0.01 ‘*’ 0.05 ‘.’ 0.1 ‘ ’ 1

Random effects:
 Groups            Name                            Variance  Std.Dev.
 country:replicate (Intercept)                     5.169e-03 0.0718951
 replicate         (Intercept)                     7.657e-03 0.0875027
 Xr.8              s(tmrca):countryIceland         1.616e-08 0.0001271
 Xr.7              s(tmrca):countryFinland         1.725e-01 0.4152843
 Xr.6              s(tmrca):countrySweden          7.605e+01 8.7208801
 Xr.5              s(tmrca):countryDenmark         2.300e+01 4.7957742
```

```
 Xr.4              s(tmrca):countryNorway       2.851e-08 0.0001688
 Xr.3              s(num_chains):countryIceland 5.111e+00 2.2606770
 Xr.2              s(num_chains):countryFinland 1.354e+00 1.1638071
 Xr.1              s(num_chains):countrySweden  3.480e+00 1.8654601
 Xr.0              s(num_chains):countryDenmark 1.077e+01 3.2814432
 Xr                s(num_chains):countryNorway  4.445e-01 0.6666932
 Residual                                       7.978e-02 0.2824595
Number of obs: 1939, groups:
country:replicate, 45; replicate, 10; Xr.8, 8; Xr.7, 8; Xr.6, 8; Xr.5, 8;
Xr.4, 8; Xr.3, 8; Xr.2, 8; Xr.1, 8; Xr.0, 8; Xr, 8
```

<u>Model: Transmission cluster duration versus number of samples</u>
(interaction with country)

```
mod.gam<-gamm4(duration ~ num_samples:country + s(num_chains, by=country)
+s(tmrca, by=country), random =~ (1|replicate/country), data=myfrm2)

Scaled residuals:
    Min      1Q  Median      3Q     Max
-5.2350 -0.3813 -0.0781  0.2943  5.2338

Parametric coefficients:
                            Estimate Std. Error t value Pr(>|t|)
(Intercept)                1.753e-01  3.632e-02   4.826 1.50e-06 ***
num_samples:countrySweden  2.256e-03  9.172e-05  24.598  < 2e-16 ***
num_samples:countryDenmark 3.637e-03  2.532e-04  14.366  < 2e-16 ***
num_samples:countryNorway  3.075e-03  4.680e-04   6.570 6.47e-11 ***
num_samples:countryFinland 4.053e-03  4.942e-04   8.202 4.31e-16 ***
num_samples:countryIceland 5.104e-03  2.163e-03   2.360   0.0184 *
---
Signif. codes:  0 ‘***’ 0.001 ‘**’ 0.01 ‘*’ 0.05 ‘.’ 0.1 ‘ ’ 1

Approximate significance of smooth terms:
                               edf Ref.df      F p-value
s(num_chains):countrySweden  7.360  7.360 62.223 < 2e-16 ***
s(num_chains):countryDenmark 4.125  4.125 25.995 < 2e-16 ***
s(num_chains):countryNorway  2.648  2.648  4.289 0.00515 **
s(num_chains):countryFinland 1.000  1.000  8.715 0.00319 **
s(num_chains):countryIceland 1.000  1.000  9.032 0.00268 **
s(tmrca):countrySweden       7.610  7.610 35.625 < 2e-16 ***
s(tmrca):countryDenmark      7.109  7.109 17.377 < 2e-16 ***
s(tmrca):countryNorway       1.000  1.000  2.697 0.10070
s(tmrca):countryFinland      2.604  2.604  2.724 0.09463 .
s(tmrca):countryIceland      1.000  1.000  0.063 0.80184
---
Signif. codes:  0 ‘***’ 0.001 ‘**’ 0.01 ‘*’ 0.05 ‘.’ 0.1 ‘ ’ 1

R-sq.(adj) =   0.79
```

```
lmer.REML = 188.32  Scale est. = 0.054151  n = 1939

Random effects:
 Groups            Name                         Variance  Std.Dev.
 country:replicate (Intercept)                  1.779e-02 1.334e-01
 replicate         (Intercept)                  4.936e-03 7.026e-02
 Xr.8              s(tmrca):countryIceland      2.704e-07 5.200e-04
 Xr.7              s(tmrca):countryFinland      7.121e-02 2.669e-01
 Xr.6              s(tmrca):countryNorway       3.274e-09 5.722e-05
 Xr.5              s(tmrca):countryDenmark      8.190e+00 2.862e+00
 Xr.4              s(tmrca):countrySweden       6.982e+01 8.356e+00
 Xr.3              s(num_chains):countryIceland 1.590e-02 1.261e-01
 Xr.2              s(num_chains):countryFinland 3.918e-06 1.979e-03
 Xr.1              s(num_chains):countryNorway  5.248e+00 2.291e+00
 Xr.0              s(num_chains):countryDenmark 2.556e+01 5.056e+00
 Xr                s(num_chains):countrySweden  6.858e+00 2.619e+00
 Residual                                       5.415e-02 2.327e-01
Number of obs: 1939, groups:
country:replicate, 45; replicate, 10; Xr.8, 8; Xr.7, 8; Xr.6, 8; Xr.5, 8;
Xr.4, 8; Xr.3, 8; Xr.2, 8; Xr.1, 8; Xr.0, 8; Xr, 8
```
